# Household transmission of SARS-CoV-2; a prospective longitudinal study showing higher viral load and transmissibility of the Alpha variant compared to previous strains

**DOI:** 10.1101/2021.08.15.21261478

**Authors:** Cathinka Halle Julin, Anna Hayman Robertson, Olav Hungnes, Gro Tunheim, Terese Bekkevold, Ida Laake, Idunn Forland Aune, Rikard Rykkvin, Dagny Haug Dorenberg, Kathrine Stene-Johansen, Einar Sverre Berg, Johanna Eva Bodin, Fredrik Oftung, Anneke Steens, Lisbeth Meyer Næss

## Abstract

**Objectives:** We studied the secondary attack rate (SAR), risk factors, and precautionary practices of household transmission in a prospective, longitudinal study. We further compared transmission between the Alpha (B.1.1.7) variant and non-Variant of Concern (non-VOC) viruses.

**Methods:** We recruited households of 70 confirmed COVID-19 cases with 146 household contacts from May 2020 to May 2021. Participants donated biological samples 8 times over 6 weeks and answered questionnaires. Whole genome sequencing and droplet digital PCR were used to establish the SARS-CoV-2 variant and viral load.

**Results:** SARS-CoV-2 transmission occurred in 60% of the households, and the overall SAR for household contacts was 50%. The SAR was significantly higher for the Alpha variant (78%) compared with non-VOC viruses (43%) and was associated with a higher viral load. SAR was higher in household contacts aged ≥40 years (69%) than in younger contacts (40-47%), and for contacts of cases with loss of taste/smell. Children had lower viral loads and were more often asymptomatic than adults. Sleeping separately from the primary case reduced the risk of transmission.

**Conclusions:** We found substantial household transmission, particularly for the Alpha variant. Precautionary practices seem to reduce SAR, but preventing household transmission may become difficult with more contagious variants.

## INTRODUCTION

SARS-CoV-2, the virus that causes the respiratory disease COVID-19, was first detected in China in 2019 and spread rapidly throughout the world. In March 2020, the World Health Organization (WHO) declared COVID-19 a pandemic. Households have been one of the most important sites of transmission in Norway (1), as well as in other countries (2). It is therefore important to identify risk factors for household transmission and effective precautionary practices to contain the epidemic. Moreover, secondary attack rate (SAR) calculated from household studies provides an important measure of the transmissibility of SARS-CoV-2.

Previous household transmission studies have mainly described transmission of the SARS-CoV-2 variants dominating in the early phase of the pandemic or have not described the genetic variant(s). However, the Alpha variant/Variant of Concern (VOC) 202012/01 (Pango lineage B.1.1.7) rapidly outcompeted other SARS-CoV-2 lineages in the UK after its emergence in November 2020 (3, 4). The first confirmed case of the Alpha variant in Norway was reported in December 2020, and from mid-February 2021 until July 2021 it was the dominant variant (1, 5). Even though increased transmissibility of the Alpha variant has been shown (6-8), knowledge is still sparse regarding how it affects the SAR in households.

We conducted a prospective longitudinal household study to investigate the SAR in Norwegian households, and to identify risk factors for transmission within these households, using frequent testing and biological sampling, together with questionnaire data. Close follow-up and systematic data collection allowed for determination of the role of viral load in transmission. We used the droplet digital PCR (ddPCR) technique to quantify SARS-CoV-2 viral RNA due to its greater accuracy and precision compared to traditional quantitative PCR (RT-qPCR) (9, 10). Moreover, we compared the SAR for the Alpha variant with the SAR for other circulating variants in Norway during the study period.

## MATERIALS AND METHODS

### Study design and study population

The design of this prospective longitudinal study was based on the WHO Household Transmission Investigation protocol (11). From May to June 2020, and from September 2020 to May 2021, we recruited households of laboratory confirmed COVID-19 cases in the capital/county Oslo and the surrounding county Viken. The course of the pandemic in Oslo/Viken, and of recruitment in this period, are shown in Figure 1A and Figure 1B, respectively. All households with a PCR-confirmed SARS-CoV-2 case aged ≥12 years, living with at least one other person aged ≥2 years, were eligible for participation. To avoid recruitment of households with co-primary cases, households with more than two members who tested positive on the same date were not eligible, unless the transmission dynamics were known. A further exclusion criterion was added when COVID-19 vaccines became available, whereby households with vaccinated individuals were not eligible.

**Figure 1:**
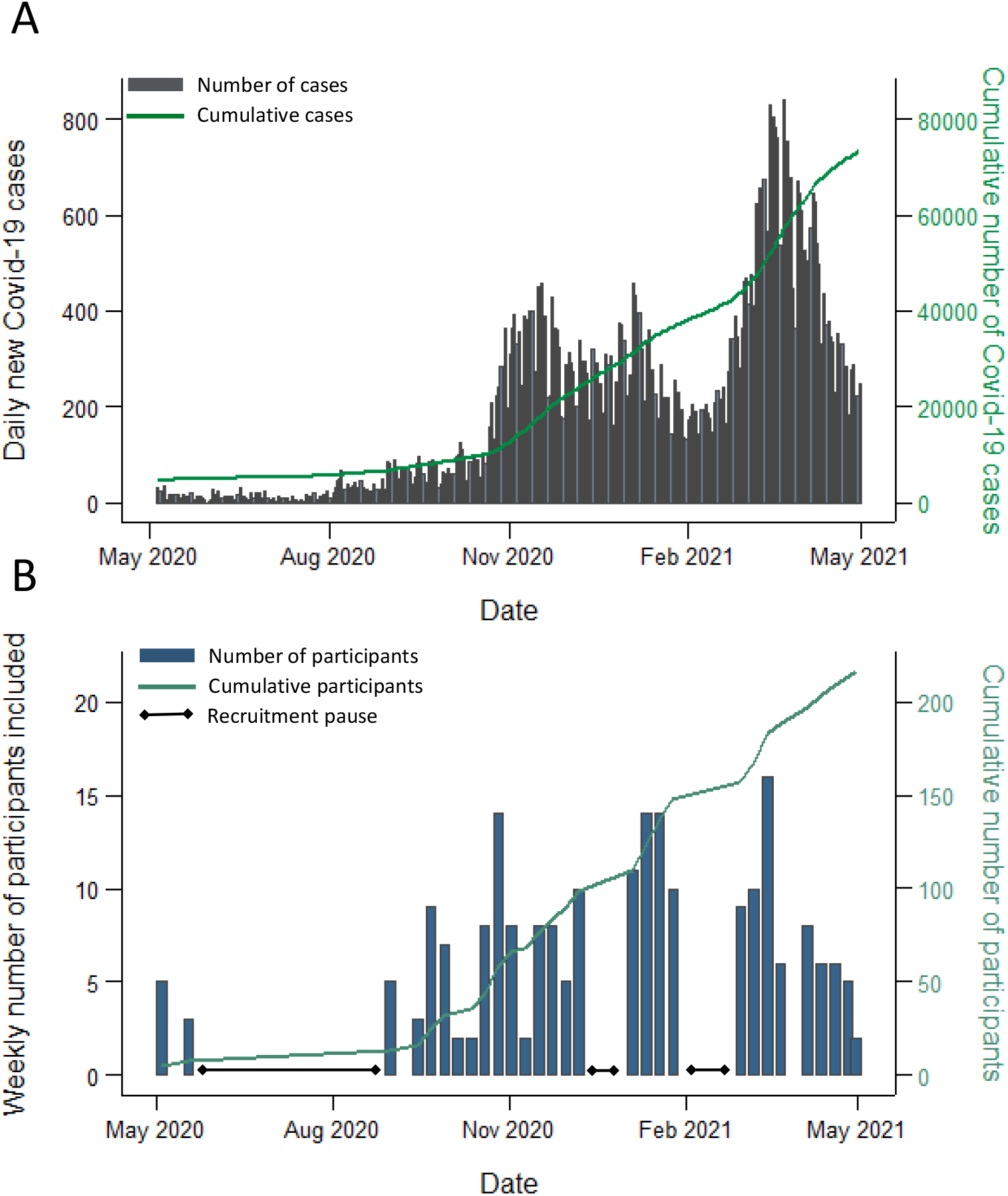
The course of the COVID-19 pandemic in Oslo/Viken, Norway, and the number of participants included in the study, from May 1 2020 to April 3 2021. A) The number of all reported laboratory confirmed COVID-19 cases in Oslo/Viken, Norway, during the study recruitment period. Participating households were recruited from these two counties. The daily number of cases is shown in grey, while the cumulative number of cases is shown in green. Source: Norwegian Surveillance System for Communicable Diseases (MSIS). B) The number of included participants during the study recruitment period. The weekly number of participants included is shown in blue, while the cumulative number is shown in green. Three recruitment pauses are indicated.

Primary cases and their household contacts were identified by the municipalities’ infection control teams following a positive SARS-CoV-2 PCR test and were subsequently contacted by the study team. Households willing to participate were visited at home, and written informed consent was obtained from the participants and/or their guardians before study inclusion.

The study was approved by the Regional Ethics Committee in Norway (#118354).

### National COVID-19 isolation and quarantine regulations

According to the Norwegian COVID-19 regulations, isolation was mandatory for persons with confirmed COVID-19. The isolation should be implemented at home or similar for at least 8-10 days after symptom debut (recommendations varied throughout the study period), lasting at least three days after symptom relief. Asymptomatic cases had to isolate for 10 days after their initial positive PCR-test. In isolation, positive cases were instructed to stay ≥2 meters from other household members, use separate bathrooms, towels, and bedrooms if possible. Household contacts were instructed to quarantine in their homes, maintaining an increased distance to other adults in the household.

### Sampling and data collection

The first home visit for inclusion and sampling was termed Day0, and seven further home visits for sampling were performed during the following 6 weeks (i.e. termed Day3, Day7, Day10, Day14, Day21, Day28, and Day42) (Supplementary Figure 1).

Oropharyngeal (OP) samples and neat saliva samples were gathered from eligible participants on each visit to test for SARS-CoV-2 by RT-qPCR. Health care workers collected OP samples using OP flocked swabs (FLOQSwabs™Copan, Italy), in 3 ml UTM (Universal Transport Medium, Copan Italy). Whole blood (Vacuette®EDTA-k2) was collected once for each participant aged ≥18 years for blood typing. Saliva and blood for immunological analyses were also collected at Day0, Day7, Day14, Day28, Day42 and Day180 (not subject of this publication).

All participants were asked to answer a questionnaire on Day0 (Q-D0), to obtain information about the household in general, transmission risk factors, clinical symptoms, and general health status. This questionnaire was adapted from the WHO protocol. The questions on behavioral risk factors in the Q-D0 related to the period up to 10 days prior to SARS-CoV-2 confirmation of the primary case, and precautionary practices after confirmation. An additional questionnaire (Q-DX) asking about the suspected source of transmission, adherence to isolation/quarantine regulations and self-report of the severity of disease, was answered by participants at the home visit on Day28/Day42 or collected through phone interviews. In addition, a symptom diary adapted from the WHO protocol was completed daily from Day0 to Day28 by all participants.

### Laboratory testing

The laboratory analysis and interpretations were conducted by NIPH. All OP and saliva samples were tested for the presence of SARS-CoV-2 by RT-qPCR. The positive samples were further analyzed by amplicon-based whole genome sequencing (WGS) of SARS-CoV-2 and droplet digital PCR (ddPCR) for absolute quantification, as described in Supplementary Methods. To estimate the viral load, SARS-CoV-2 RNA copies per µl eluate was determined by ddPCR using the saliva sample with the lowest cycle of threshold (Ct) value for each participant, if sufficient material was available.

### Definition of cases and contacts

Household contacts were defined as individuals aged ≥2 years who resided with the primary case. A household contact was considered a secondary case if they had a positive PCR test (OP and/or saliva), and their symptom onset/PCR positive test (which ever came first; defined as T0) was within 14 days after T0 of the primary case. If a household contact had a T0 ≥2 days prior to T0 of the original primary case, the household contact was defined as an alternative primary case. If household members had the same T0, or +/-1 day, they were re-defined as co-primary cases, unless the original primary case had a known source of infection outside of the household.

### Study samples included in analysis

For the main overall SAR analysis, households containing co-primary cases were excluded. Households with alternative primary cases were included in the overall SAR analysis but excluded from the analysis on behavioral factors and preventive measures due to lack of data from the Q-D0 questionnaire (Figure 2). For comparisons between genetic variants, households with the Alpha lineages were compared with non-VOC SARS-CoV-2 viruses, hereby referred to as non-VOC viruses (12), while households with other VOCs were excluded from the analyses (i.e. one household with the Beta variant). One household contact lacked variant data and was assigned the same variant as the primary case.

**Figure 2:**
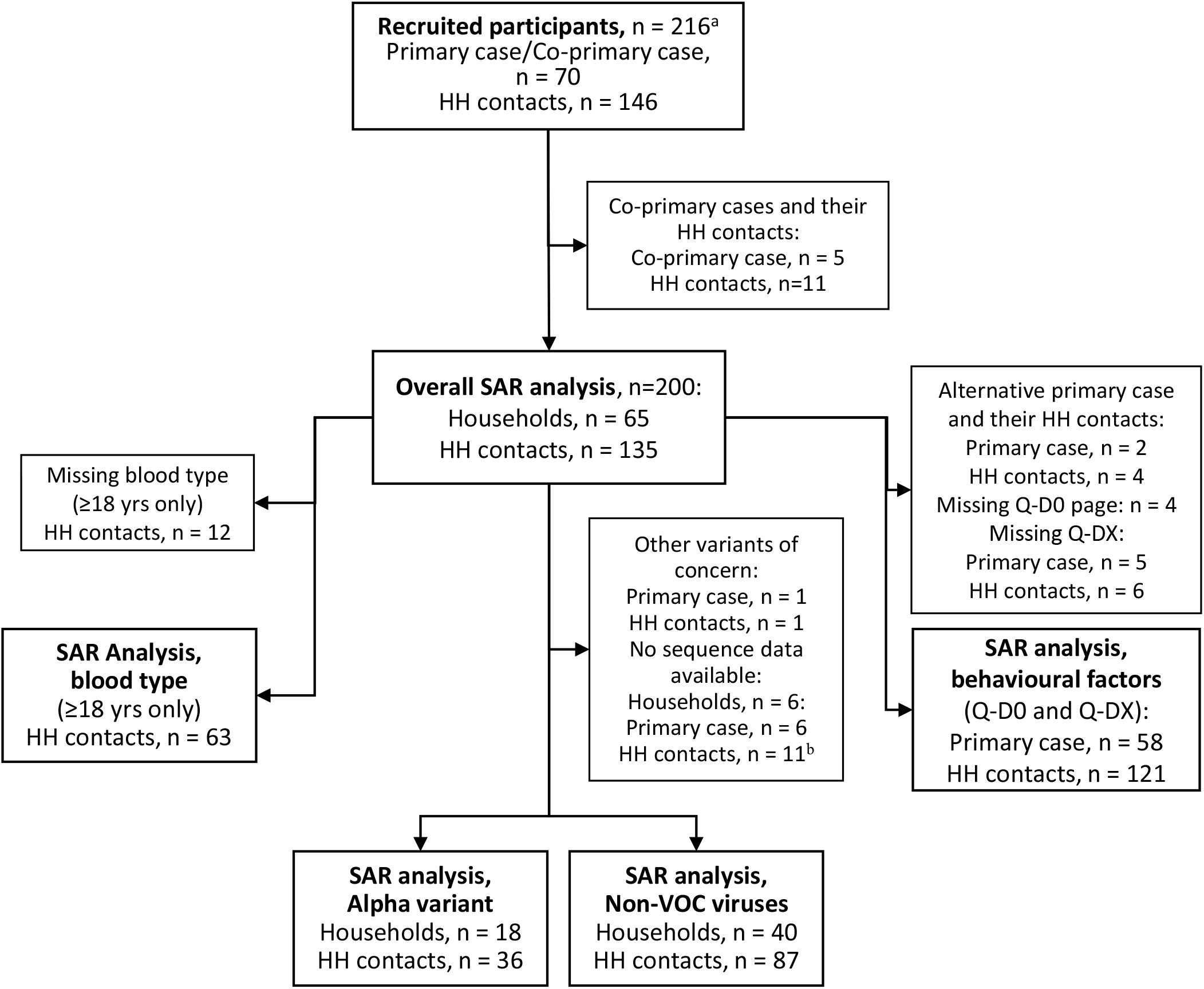
Flow chart of participant selection for the different analyses. Abbreviations: HH contact, household contact; non-VOC, non-Variant of Concern ^a^ recruited participants were tested for SARS-CoV-2 by RT-qPCR and provided information on symptom onset (Q-D0 questionnaire) ^b^ includes household contacts that were SARS-CoV-2 negative

### Data analysis and statistical methods

The SAR was estimated as the proportion (%) of household contacts that were defined as confirmed cases (11). Cluster robust standard errors were used to calculate 95% confidence intervals. The proportion of households with secondary transmission was also estimated. To test for differences in proportions, the Pearson chi-square test statistics was corrected with the second-order correction of Rao and Scott and converted into an F statistic (13).

To account for dependencies within households, a mixed-effect logistic regression model with a household-level random intercept was used to study the associations between potential risk factors for transmission and of infection among the household contacts. The multivariable models were adjusted for age and sex of the household contacts and of the primary cases, and household size. The analysis on associations between SARS-CoV-2 viral load (SARS-CoV-2 RNA copies/ul eluate measured by ddPCR) and symptoms was limited to the confirmed cases. For analyses with all cases, a mixed-effect logistic regression adjusted for age and sex was used, whereas for analyses only done on primary cases logistic regression was used. To study the association between genetic variant and viral load, a mixed-effect linear regression adjusted for age and sex was used. The mean duration of detectable SARS-CoV-2 was estimated for household contacts only, as described in Supplementary Methods for children (<18 years) and adults (≥ 18 years), and cluster robust standard errors were used to calculate 95% confidence intervals. Primary cases were not included in this analysis as the majority were adults (due to the inclusion criteria of the study) and infection was likely detected later in the course of disease for these participants. To estimate the association between duration of detectable SARS-CoV-2 by RT-qPCR (in days) and age group, a mixed-effect linear regression was used.

All analyses were performed in STATA/SE 15.0 (StataCorp. College Station, Texas USA). A p-value of <0.05 was considered statistically significant (shown in bold in the tables).

## RESULTS

### Baseline characteristics of households and participants

We recruited 70 households, including 216 participants (Figure 2). Ninety eight percent of eligible household members agreed to participate in the study. Five cases were co-primary cases and were excluded together with their 11 household contacts. A total of 65 primary cases/households and their 135 household contacts (200 participants) were thus eligible for the evaluation of secondary transmission. Among the 65 households, 18 of the primary cases were infected with the Alpha variant, one with the Beta variant and 40 with other circulating non-VOC viruses (Supplementary Table S1). Households with the Alpha variant were recruited between March and May 2021, while households with non-VOC viruses were mainly recruited before February 2021, reflecting the viral circulation in the study area during the recruitment period (Supplementary Figure 2). Sequence data showed the same genetic lineage for all sequenced members within individual households. Demographic and clinical characteristics of the participants in the SAR analyses are shown in Table 1. The median age of the participants was 31 years, and primary cases were generally older than household contacts (38 and 24 years, respectively). About 1/3^rd^ of the participants were children aged <18 years, while only six were older than 65. The proportion of males and females was equal, and 51% were of Nordic ethnicity (but there was considerable missing data for this variable).

**Table 1.**
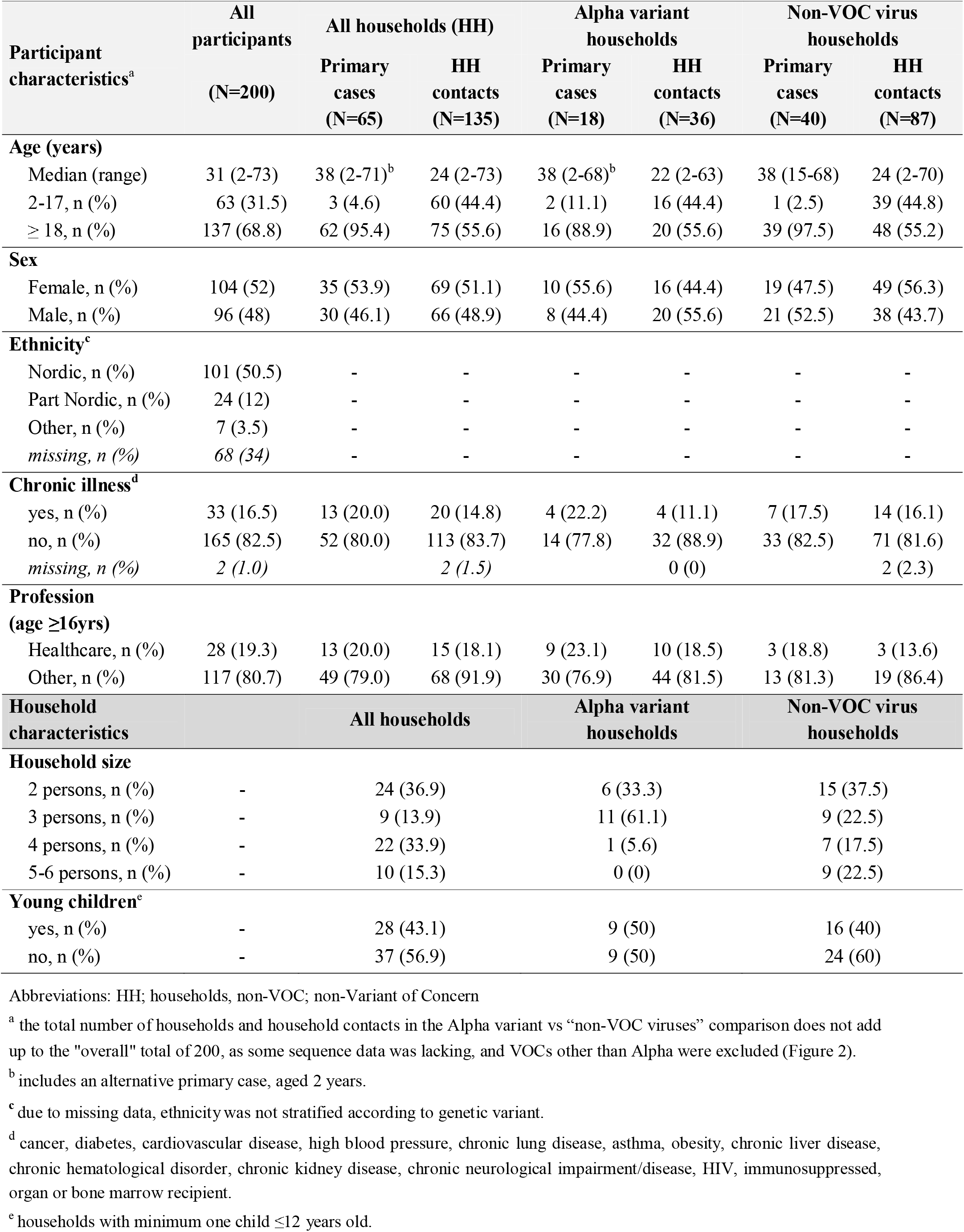
Demographic and clinical characteristics of all included participants and households, and Alpha variant and non-VOC virus households

The median household size was four, ranging from two to six people, and families with young children constituted 43% of the households (Table 1). The household size was slightly smaller in households where participants were infected with the Alpha genetic variant (median = 3), compared with households infected with non-VOC viruses (median = 4). The remaining characteristics were broadly similar between these two groups.

Of the 200 participants 132 (66%) were infected. Fourteen percent of the confirmed cases were asymptomatic, while 43% had mild disease and 42% had a moderate disease, based on their reported symptoms within 14 days of their first positive PCR sample. Few study participants were hospitalized, and all were discharged the following day. There were slightly more asymptomatic cases (22%) among the Alpha variant participants compared with participants with non-VOC viruses (9%), although the difference was not significant (p=0.09) (Supplementary Table S2). Severity also varied with age, with 36% of children (<18 years) being asymptomatic compared to 8% of adults (p<0.01). Children were SARS-CoV-2 RT-qPCR positive for a shorter time period than adults (mean number of days 11.3 (95% CI 7.6-15.1) and 16.4 (95% CI 13.5-19.3) respectively, p=0.03).

### Secondary transmission of COVID-19 in households

Secondary transmission occurred in 60.0 % of the households in the study (95% CI 47.4-71.4) (Table 2). The secondary attack rate (SAR) among all household contacts was 49.6% (95% CI 37.8–61.5). Secondary transmission was significantly higher in households with the Alpha variant (83.3%, 95% CI 55.9–95.2) compared with non-VOC viruses (55.0% (95% CI 39.8–70.1), p=0.04). For household contacts, SAR was 77.8% (95% CI 49.4-92.6) in households with the Alpha variant, compared with 42.5% (95% CI 28.7-57.7) in households with non-VOC viruses, resulting in a significantly higher adjusted odds ratio (OR) for secondary transmission in households with the Alpha variant (p=0.03) (Table 2).

**Table 2.**
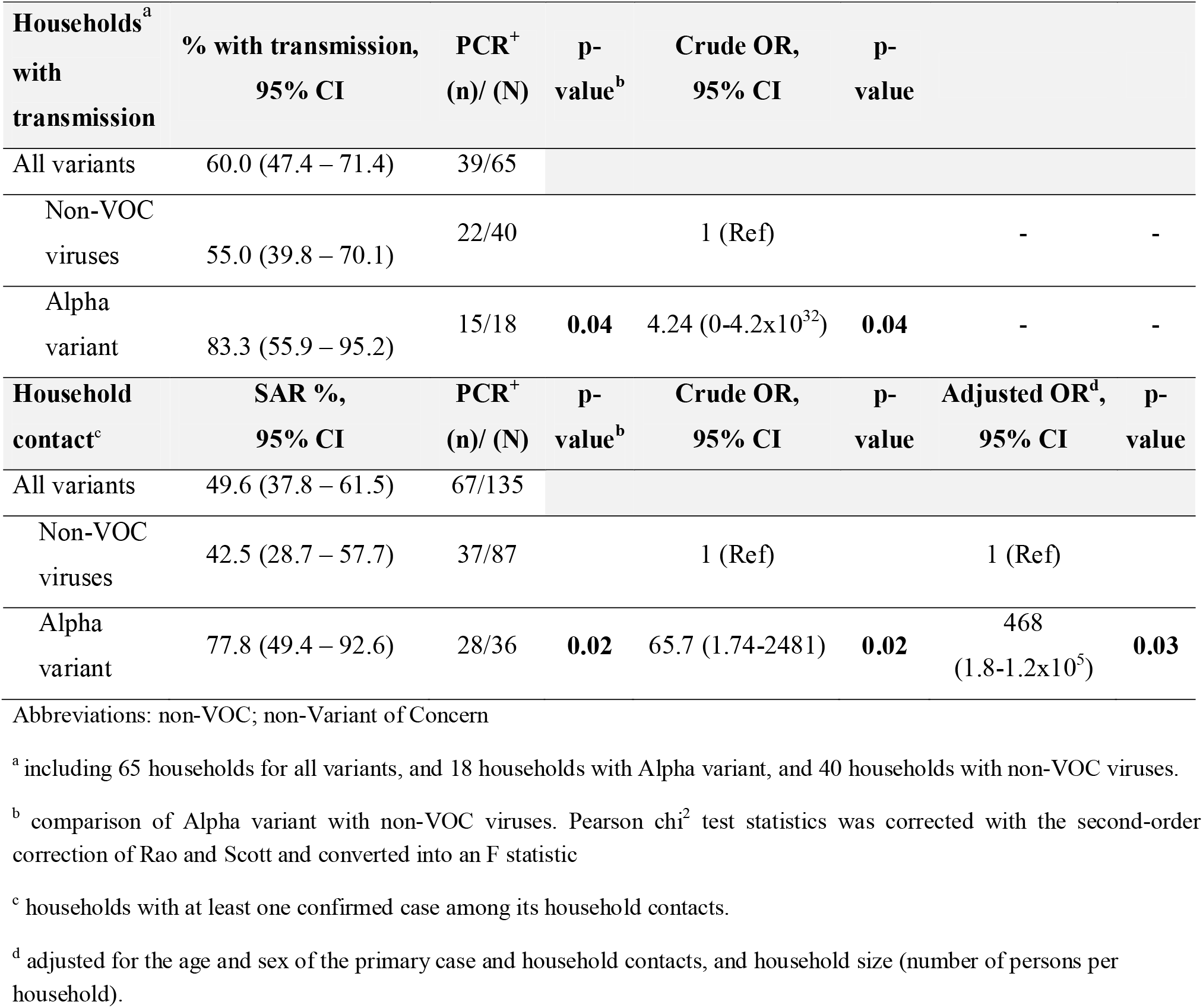
Comparison of transmission rates for all households and household contacts, and for Alpha variant versus non-VOC viruses

The median interval from the date of the first positive SARS-CoV-2 test (collected by the municipality for the primary case) and the Day0 visit in the study was 3 days (IQR; 2-4 days). A large proportion of the secondary cases (38.5%) were already infected at Day0, while 61.5% of the secondary cases were detected during study follow-up. The overall serial interval (the number of days between symptom onset of the primary case and of a household contact) was estimated to 4 days (range 1-11, n=50). The median serial interval was similar for the Alpha variant (4 days, range 2-11, n=17) and non-VOC viruses (4 days, range 1-9 days, n=31). The overall median interval between symptom onset of the primary case and the first RT-qPCR-positive test of a household contact was 3 days (range 1-12, n=60), and this interval was the same for Alpha (3 days, range 1-11, n=25) and non-VOC viruses (4 days, range 1-9 days, n=33).

### Effect of host and household characteristics on secondary transmission

Neither age (12-39 years compared to ≥40 years) nor sex of the primary case appeared to have an impact on SAR (Table 3). Notably there were few primary cases under the age of 18, therefore it was not possible to study the effect of age on transmission for primary cases aged 12-18 years.

**Table 3.** Secondary attack rates (SAR) and odds ratios (OR) for secondary infection of all household contacts (N=135) according to characteristics of primary case, household contact characteristics, and household characteristics.

| Characteristic | SAR %, (95% CI) | p-value<sup>a</sup> | PCR<sup>+</sup> (n) / total (N) | Crude OR (95% CI) | p-value | Adjusted OR<sup>b</sup> (95% CI) | p-value |
| --- | --- | --- | --- | --- | --- | --- | --- |
| **PRIMARY CASE CHARACTERISTICS** |  |  |  |  |  |  |  |
| **Age (yrs)** |  |  |  |  |  |  |  |
| 12-39<sup>c</sup> | 47 (31-64) |  | 33/70 | 1 (Ref) |  | 1 (Ref) |  |
| ≥40 | 52 (35-69) | 0.67 | 34/65 | 2.30 (0.25- 21.1) | 0.46 | 1.6 (0.15- 16.9) | 0.70 |
| **Sex** |  |  |  |  |  |  |  |
| Female | 46 (30-63) |  | 29/63 | 1 (Ref) |  | 1 (Ref) |  |
| Male | 53 (35-70) | 0.58 | 38/72 | 1.51 (0.18- 12.7) | 0.71 | 1.7 (0.15- 18.7) | 0.68 |
| **HOUSEHOLD CONTACT CHARACTERISTICS** |  |  |  |  |  |  |  |
| **Age (yrs)** |  |  |  |  |  |  |  |
| 2-17yr | 47 (31-63) |  | 28/60 | 1 (Ref) |  | 1 (Ref) |  |
| 18-39 | 40 (25-56) |  | 17/43 | 0.31 (0.05-2.04) | 0.23 | 0.18 (0.02-1.33) | 0.09 |
| ≥40 | 69 (49-83) | **0.03** | 22/32 | 8.03 (1.15- 56.2) | **0.04** | 7.53 (1.07-52.8) | **0.04** |
| **Sex** |  |  |  |  |  |  |  |
| Female | 52 (38-66) |  | 36/69 | 1 (Ref) |  | 1 (Ref) |  |
| Male | 47 (32-62) | 0.55 | 31/66 | 0.81 (0.22- 3.01) | 0.76 | 0.97 (0.24- 3.91) | 0.96 |
| **Blood type (≥18yrs)** |  |  |  |  |  |  |  |
| O | 48 (27-69) |  | 10/21 | 1 (Ref) |  | 1 (Ref) |  |
| A | 56 (37-73) |  | 18/32 | 1.45 (0.38- 5.5) | 0.59 | 1.41 (0.40- 4.96) | 0.59 |
| AB | 33 (0-100) |  | 1/3 | 0.49 (0.02-10.3) | 0.64 | 0.40 (0.02- 6.47) | 0.52 |
| B | 71 (22-96) | 0.61 | 5/7 | 3.03 (0.32-28.3) | 0.33 | 3.02 (0.41- 22.5) | 0.28 |
| **HOUSEHOLD CHARACTERISTICS** |  |  |  |  |  |  |  |
| **Household size** |  |  |  |  |  |  |  |
| 2 pers | 54 (33-74) |  | 13/24 | 1 (Ref) |  | 1 (Ref) |  |
| 3 pers | 59 (27-84) |  | 10/17 | 1.72 (0.06- 53.0) | 0.76 | 2.0 (0.05- 88) | 0.71 |
| 4 pers | 47 (28-68) |  | 28/59 | 0.48 (0.03- 7.72) | 0.61 | 0.8 (0.04- 17) | 0.88 |
| 5-6 pers | 46 (20-74) | 0.87 | 16/35 | 0.48 (0.15- 18.6) | 0.68 | 0.7 (0.02- 29) | 0.84 |
| **Overcrowding<sup>d</sup>** |  |  |  |  |  |  |  |
| No | 52 (37-66) |  | 47/91 | 1 (Ref) |  | 1 (Ref) |  |
| Yes | 90 (24-100) | **0.01** | 9/10 | 122.7 (0.16- 94464) | 0.16 | 480.9 (0.11-2x10<sup>6</sup>) | 0.15 |
| **Number of bathrooms<sup>e</sup>** |  |  |  |  |  |  |  |
| 1 | 58 (39-75) |  | 36/62 | 1 (Ref) |  | 1 (Ref) |  |
| ≥2 | 44 (27-61) | 0.25 | 27/62 | 0.2 (0.02- 2.7) | 0.25 | 0.1 (0.01- 2.2) | 0.17 |

Abbreviations: SAR; Secondary Attack Rate, OR; Odds Ratio, CI; Confidence Interval
<sup>a</sup> Pearson chi<sup>2</sup> test statistics was corrected with the second-order correction of Rao and Scott and converted into an F statistic
<sup>b</sup> adjusted for the age and sex of the primary case and household contacts, and household size (number of persons per household), unless this was the factor being analyzed.
<sup>c</sup> includes an alternative primary case, age 2
<sup>d</sup> data missing for 34 household contacts (25%). Overcrowding was defined as 1) the number of rooms in the property being lower than the number of persons living in the household, AND 2) the number of square meters being less than 25 per person.
<sup>e</sup> data missing for 21 household contacts (16%)

Secondary infection amongst children aged 2-17 years was similar for those aged 18-39 (SAR 47% and 40%, respectively), while household contacts aged ≥40 years were more likely to be infected (69%) (Table 3). The sex and blood type of the household contacts did not impact the infection risk. Household contacts living in overcrowded houses had a higher infection risk than those not living in overcrowded houses (SAR 90% and 52%, respectively), but the difference was not significant when adjusted for age, sex, and household size. However, the number of overcrowded households was small. Secondary transmission did not differ with household size or number of bathrooms in the household.

Both fever and loss of taste/smell were significantly more common in primary cases with the Alpha variant compared to others (Supplementary Table S3). In addition, the SAR was higher if these symptoms were present (Table 4). If the primary case reported loss of taste/smell, the SAR was 60% versus 27%, and there was a similar trend for fever (61% versus 39%), and a weak trend for cough. Dyspnea in the primary case did not appear to influence the SAR, nor clinical severity.

**Table 4.** Secondary attack rates (SAR) and odds ratios (OR) for secondary infection for all household contacts (N=135) according to clinical severity and symptoms of primary case.

|  | SAR %<br>(95%CI) | PCR<sup>+</sup><br>(n)/<br>total (N) | p-<br>value<sup>a</sup> | Crude OR<br>(95%CI) | p-<br>value | Adjusted<sup>b</sup> OR<br>(95%CI) | p-<br>value |
| --- | --- | --- | --- | --- | --- | --- | --- |
| **Severity** |  |  |  |  |  |  |  |
| Asymptomatic | 33 (3-88) | 4/12 |  | 1 (Ref) |  | 1 (Ref) |  |
| Mild | 47 (27-68) | 25/53 |  | 10.2 (0.17-613) | 0.27 | 8.7 (0.13-594) | 0.31 |
| Moderate | 54 (39-69) | 38/70 | 0.54 | 12.1 (0.22-672) | 0.22 | 11.8 (0.14-974) | 0.28 |
| **Loss of taste/ smell** |  |  |  |  |  |  |  |
| No | 27 (13-47) | 12/44 |  | 1 (Ref) |  | 1 (Ref) |  |
| Yes | 60 (45-74) | 55/91 | <0.01 | 29.5 (1.33-654) | 0.03 | 68.3 (1.95-2389) | 0.02 |
| **Fever** |  |  |  |  |  |  |  |
| No | 39 (24-57) | 27/69 |  | 1 (Ref) |  | 1 (Ref) |  |
| Yes | 61 (43-76) | 40/66 | 0.08 | 10.3 (0.78-136) | 0.08 | 10.4 (0.76-140) | 0.08 |
| **Cough** |  |  |  |  |  |  |  |
| No | 27 (11-53) | 7/26 |  | 1 (Ref) |  | 1 (Ref) |  |
| Yes | 55 (41-68) | 60/109 | 0.04 | 10.0 (0.54-184) | 0.12 | 10.2 (0.51-203) | 0.13 |
| **Dyspnea** |  |  |  |  |  |  |  |
| No | 46 (29-64) | 31/68 |  | 1 (Ref) |  | 1 (Ref) |  |
| Yes | 54 (38-69) | 36/67 | 0.50 | 1.40 (0.17-12) | 0.76 | 0.97 (0.1-9.51) | 0.98 |

Abbreviations: SAR; Secondary Attack Rate, OR; Odds Ratio, CI; Confidence Interval
<sup>a</sup> Pearson $\chi^2$ test statistics was corrected with the second-order correction of Rao and Scott and converted into an F statistic
<sup>b</sup> adjusted for the age and sex of the primary case and household contacts, and household size (number of persons per household).

### Role of viral load measured by ddPCR

As expected, the correlation between viral load (SARS-CoV-2 RNA copies/µl eluate) determined by ddPCR and the RT-qPCR Ct-values was strong (r = -0.859, p < 0.001). There was a trend that higher viral load measured by ddPCR was associated with increased risk of secondary infection (adjusted OR 3.05 (95% CI 0.84-11.0), p=0.089). Higher viral load was also associated with increased risk of loss of taste/smell (adjusted OR = 1.4 (95% CI 1.06-1.85), p=0.02), and possibly dyspnea (OR = 1.34 (95% CI 0.96-1.86), p=0.08) and cough (adjusted OR 1.37 (95% CI 0.93-2.01), p=0.11) (Supplementary Table S4). However, despite an OR larger than 1, these associations were not significant when looking at only the primary cases, possibly because of the lower sample size.

The viral load was significantly higher for the Alpha variant than for non-VOC viruses (mean 3.24 log_10_ and 2.48 log_10_ RNA copies/µl eluate, respectively) (Figure 3A). We also found a significantly lower viral load in children than in adults (mean 2.09 log_10_ copies/µl RNA and 2.98 log_10_ copies/µl RNA, respectively) (Figure 3B), irrespective of virus variant (Figure 3C). The association between viral load and the Alpha variant remained significant in a mixed-effect linear regression model when adjusted for age and sex (adjusted regression coefficient of 0.87 (95% CI 0.34-1.40), p= 0.001).

**Figure 3:**
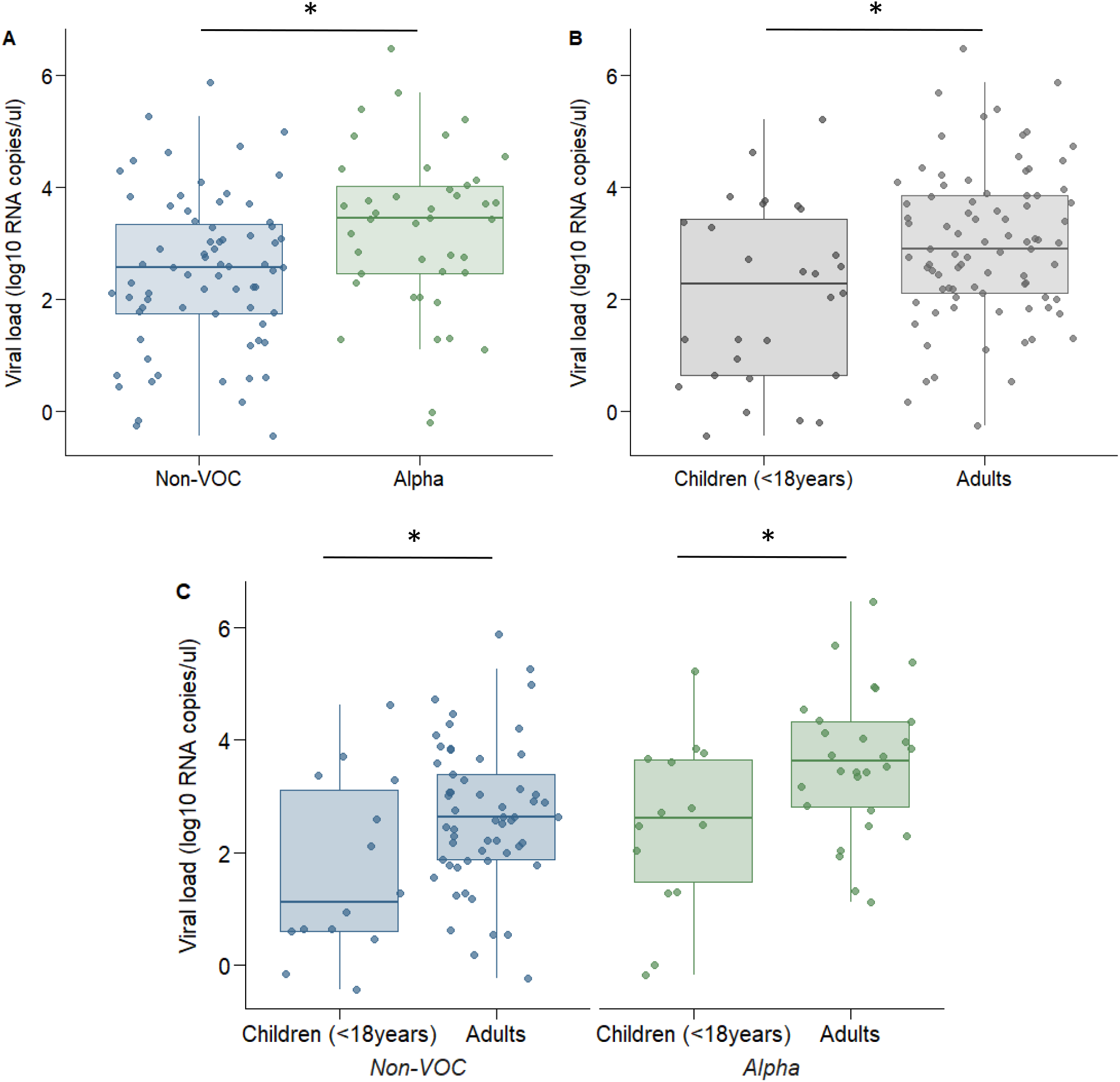
Comparison of viral load (log10 RNA copies/µl eluate) measured by ddPCR for genetic variant and age groups. Abbreviations: non-VOC, non-Variant of Concern A) Comparison of viral load between non-VOC (n=71) and Alpha viruses (n=42) B) Comparison of viral load between children (n=28) and adults (n=85) for all genetic variants combined C) Comparison of viral load for genetic variants and age groups: non-VOC; children (n=14) versus adults (n=57), and Alpha variant; children (n=14) versus adults (n=28) * p < 0.05. P-values were estimated using a mixed-effects linear regression.

### The impact of behavioral factors and precautionary practices on secondary transmission

None of the contact behaviors between the primary case and the household contacts prior to confirmation of infection of the primary case were significantly associated with SAR (Table 5). Nevertheless, there was a trend that the SAR was higher for contacts who shared a toilet, hugged, kissed, shook/ held hands, slept in the same room and shared a bed with the primary case before infection was confirmed.

**Table 5.** Effect of behavioral factors and precautionary practices on secondary attack rate (SAR).

|  |  | SAR%<br>(95% CI) | PCR<sup>+</sup>/<br>HH<br>contacts | p-<br>value <sup>a</sup> | Crude OR<br>(95% CI) | p-<br>value | Adjusted OR<sup>b</sup><br>(95% CI) | p-<br>value |
| --- | --- | --- | --- | --- | --- | --- | --- | --- |
| **Behavioral factors: contact with the primary case *prior* to confirmation of infection** |  |  |  |  |  |  |  |  |
| Cared for | No | 48 (35-61) | 51/106 |  | 1 (Ref) |  | 1 (Ref) |  |
|  | Yes | 53 (27-78) | 8/15 | 0.70 | 1.14 (0.31-2.74) | 0.91 | 0.54 (0.04–6.79) | 0.63 |
| Hugged | No | 39 (21-61) | 11/28 |  | 1 (Ref) |  | 1 (Ref) |  |
|  | Yes | 52 (37-66) | 48/93 | 0.28 | 3.26 (0.54-9.60) | 0.20 | 3.90 (0.55-27.7) | 0.17 |
| Kissed | No | 44 (29-60) | 26/59 |  | 1 (Ref) |  | 1 (Ref) |  |
|  | Yes | 53 (36-70) | 33/62 | 0.41 | 7.14 (0.86-9.10) | 0.07 | 5.27 (0.63-44.0) | 0.13 |
| Shook/<br>held hands | No | 47 (30-65) | 16/34 |  | 1 (Ref) |  | 1 (Ref) |  |
|  | Yes | 49 (36- 63) | 43/87 | 0.80 | 3.51 (0.56- 22.02) | 0.18 | 4.30 (0.52- 35.5) | 0.18 |
| Ate together | No | 50 (25-75) | 6/12 |  | 1 (Ref) |  | 1 (Ref) |  |
|  | Yes | 49 (36-62) | 53/109 | 0.91 | 1.58 (0.20- 2.78) | 0.67 | 1.72 (0.19- 15.1) | 0.63 |
| Shared a<br>cup/glass/bottle | No | 49 (36-62) | 50/102 |  | 1 (Ref) |  | 1 (Ref) |  |
|  | Yes | 47 (20-77) | 9/19 | 0.92 | 0.85 (0.10- 0.25) | 0.88 | 0.60 (0.06- 5.64) | 0.65 |
| Slept in the<br>same room | No | 43 (26-61) | 25/58 |  | 1 (Ref) |  | 1 (Ref) |  |
|  | Yes | 54 (39- 68) | 34/63 | 0.30 | 3.40 (0.79-4.66) | 0.10 | 2.58 (0.53- 12.5) | 0.24 |
| Shared a bed | No | 42 (27-60) | 25/59 |  | 1 (Ref) |  | 1 (Ref) |  |
|  | Yes | 55 (40-69) | 34/62 | 0.19 | 2.93 (0.68-2.54) | 0.15 | 2.18 (0.45-10.5) | 0.34 |
| Shared a toilet | No | 20 (2-71) | 2/10 |  | 1 (Ref) |  | 1 (Ref) |  |
|  | Yes | 51 (38-65) | 57/111 | 0.10 | 117 (0.5-26964) | 0.085 | 125 (0.5-29152) | 0.08 |
| **Precautionary practices: performed by primary cases *after* confirmation of infection** |  |  |  |  |  |  |  |  |
| Isolated<sup>c</sup> | No | 67 (40-86) | 33/22 |  | 1 (Ref) |  | 1 (Ref) |  |
|  | Yes | 42 (28-57) | 37/88 | 0.10 | 0.07 (0.00- 1.15) | 0.06 | 0.06 (0.00- 1.19) | 0.07 |
| Social<br>distanced<br>(≥2m) | No | 59 (40-76) | 35/59 |  | 1 (Ref) |  | 1 (Ref) |  |
|  | Yes | 39 (23-57) | 24/62 | 0.11 | 0.11 (0.01-1.31) | 0.08 | 0.10 (0.01-1.32) | 0.08 |
| Used<br>face mask | No | 55 (38-70) | 46/84 |  | 1 (Ref) |  | 1 (Ref) |  |
|  | Yes | 35 (19-56) | 13/37 | 0.12 | 0.13 (0.01- 1.62) | 0.11 | 0.12 (0.01- 1.51) | 0.10 |
| Slept in a<br>different room | No | 67 (44-85) | 29/43 |  | 1 (Ref) |  | 1 (Ref) |  |
|  | Yes | 38 (25-54) | 30/78 | **0.03** | 0.08 (0.01- 0.98) | **0.048** | 0.07 (0.00- 0.98) | **0.048** |
| Used separate<br>bathroom/ toilet | No | 53 (38-68) | 41/77 |  | 1 (Ref) |  | 1 (Ref) |  |
|  | Yes | 41 (21 -64) | 18/44 | 0.35 | 0.30 (0.03-3.00) | 0.31 | 0.30 (0.02- 3.63) | 0.34 |
| Did not share a<br>towel/ items<sup>d</sup> | No | 69 (25-94) | 11/16 |  | NA |  | NA |  |
|  | Yes | 49 (32-67) | 30/61 | 0.34 |  |  |  |  |

Abbreviations: SAR; Secondary Attack Rate, OR; Odds Ratio, CI; Confidence Interval
<sup>a</sup> Pearson chi<sup>2</sup> test statistics was corrected with the second-order correction of Rao and Scott and converted into an F statistic<sup>b</sup> adjusted for age and sex of the primary case and household contacts, and household size (number of persons per household)<sup>c</sup> defined as resided in a separate room and kept ≥2 meters distance from the rest of the household members, did not share a bedroom<sup>d</sup> if shared a bathroom/toilet

After confirmation of the infection of the primary case, the only precautionary practice to significantly prevent household transmission was sleeping in a separate room from the primary case, with a SAR of 38 %, compared to 67% for those who slept in the same room (p=0.048) (Table 5). All other precautionary practices tended to lower the SAR, particularly isolation of the primary case, but associations were not statistically significant.

## DISCUSSION

This prospective longitudinal household study with close follow-up and systematic sampling shows a high overall SAR (49.6%), confirming that households are an important site of transmission. The SAR of the Alpha variant (B.1.1.7 VOC) was significantly higher, at 77.8%, compared with 42.5% for the other non-VOC viruses dominating in Norway until Feb/March 2021. A significantly higher viral load was found in the saliva of participants with the Alpha variant compared to the non-VOC viruses, which may contribute to the increased transmissibility. Close contact behavior prior to confirmation of infection of the primary case tended to give a higher SAR. However, we showed that SAR was reduced if the primary case slept in a separate room or was isolated from the rest of the household after infection was confirmed.

Our SAR-estimate of 42.5% for non-VOC viruses is higher than the estimates of around 17 % found in other early reviews (2, 14), but is in accordance with another Norwegian household study from the first wave of the pandemic, which estimated a SAR of 47% based on RT-qPCR and seroconversion (15). Other studies performed in the UK, the Netherlands, and the US in the beginning/middle of 2020 also found similar SARs of 37%-53% (16-18). A more recent Norwegian national register-based study found a considerably lower household SAR of only 21% (19). Register based studies are more sensitive to underreporting, as it is not mandatory to test all household members, which may in turn lead to an underestimation of SAR. In particular, parents may hesitate to test children because of discomfort with nasopharyngeal swabbing. Indeed, Fung *et al*. (14) showed that studies that tested household members more frequently observed higher SARs. None of the aforementioned studies sequenced positive virus samples or quantified viral load, and most were performed before the Alpha strain appeared.

The Alpha variant has been shown to be generally more transmissible than non-VOC viruses (7, 20) and our study demonstrates this in a household setting. Our finding that SAR is significantly higher in households with the Alpha variant compared with non-VOC viruses, is in agreement with previous household studies (21-23). However, we estimated a substantially higher SAR (77.3%) for the Alpha variant than was reported in these other studies (25.1%, 38%, and 42%), probably because they were registry based. In our study, the extensive testing over several weeks with both salivary and OP samples, including testing of small children, probably enabled identification of most infected cases in the households, and thus contributed to our higher SAR estimates both overall and for the Alpha variant in particular. We found no difference between the median serial interval for the Alpha variant and the non-VOC viruses, which is in accordance with other studies (24).

Previous estimates of SAR in children and different age groups, have been conflicting (2, 18, 25-29), probably due to various biases, as discussed by Goldstein and colleagues (29). We found that the risk of transmission was similar for children (<18 years) and adults below 40 years, while household contacts aged ≥40 years had increased risk of secondary transmission. The age of the primary case was not associated with the risk of secondary transmission in the household. However, most of the primary cases in our study were >18 years old.

We used ddPCR to accurately assess SARS-CoV-2 viral load and to avoid potential inference from inhibitory substances which may influence the results when using RT-qPCR for quantification (9). Previous studies have been conflicting regarding the relationship between viral variant and viral load (21, 30, 31). Our results support that the Alpha variant is associated with a higher viral load. It has been argued that the time of sampling may obscure the comparison of viral loads between variants (30). In our study, frequent sampling enabled the selection of the sample with the lowest Ct-value for the quantification of viral load by ddPCR, thus reducing the effect of timing of sampling collection. Furthermore, this finding was consistent when the analysis was limited to the household contacts only (data not shown), for whom sampling was performed earlier in the course of infection compared with the primary cases. We also demonstrated both lower viral load and shorter duration of viral detection in children compared with adults, which in accordance with some studies (32-34), while others have shown no difference in viral load (35). Our results may suggest that it is more difficult to detect active infection in children, and that the timing of the test is of importance. Further, loss of taste/smell in primary cases, a distinctive feature of COVID-19 infection (36), was associated with a significant increase in SAR, which may in part be explained by an increased viral load as observed in participants reporting loss of taste/smell. The association between taste/smell impairment and higher viral load has also been found by others (37, 38). This may be dependent on variant, as we found that loss of taste/smell was more common amongst primary cases with the Alpha variant.

Most contact behavior such as kissing, appeared to slightly increase the odds of secondary transmission, although not significantly. We found that sleeping separately from the primary case after confirmation of infection prevented secondary infection, as shown previously (28). Other measures reducing contact with the primary case, especially isolation, also seemed to lower secondary transmission. This is in contrast to a similar household study by Miller and colleagues (25) which found no effect, possibly explained by transmission already occurring prior to laboratory confirmation of the primary case. Although we also observed that a high fraction of the transmission had occurred quite early, our findings still support the importance of starting precautionary practices after infection.

The present study has several limitations. First, our sample size was small, which limited the comparison between factors associated with the Alpha variant and other non-VOC viruses. Further, the study was not initially designed to evaluate differences in SAR between variants, and the dominance of the variants differed during the study period. We can therefore not exclude that climate, people’s behavior, or other factors, could have influenced our results. Quarantine and isolation guidelines were similar throughout the whole study period; thus, we assume that this has not significantly influenced our results. Finally, the age span of participants was limited, with few elderly individuals and mostly adult primary cases.

In conclusion, in this prospective longitudinal household study, we found an overall secondary attack rate for household contacts of 49.6%. The SAR was considerably higher for the Alpha variant (77.8%) than for non-VOC viruses, indicating a very high level of household transmission, possibly mediated by the higher viral load, for this VOC. We also showed that age affects secondary infection, with higher SAR in household contacts older than 40 years. Implementation of precautionary measures after detection of the first SARS-CoV-2 case seems to reduce household transmission, in particular sleeping separately from the primary case. However, preventing transmission within a household will become increasingly difficult with the emergence of more contagious variants. Our results emphasize the role of households in the transmission of SARS-CoV-2 in the Norwegian population and the importance of strict adherence to the isolation and quarantine regulations in all households with a confirmed case.

## Data Availability

Data are not publicly available

## Acknowledgements

We want to thank all the participants who willingly let us into their homes and provided frequent samples over a long period of time. We also want to acknowledge the following; our eminent health care workers who visited and sampled all the participants (Marit F. Killengren, Torunn R. Strand, Hena Anawar, Christina Nitschke, Kristina Maudal); the head of Section for Influenza (Karoline Bragstad); the coordinators, technicians and bioinformaticians at the Department of Virology, the Department of Bacteriology, the Reception of Biological Samples; the engineers at the Department of Method Development and Analytics; the municipalities’ infection control teams in Oslo/Viken for their contribution to recruitment of participants; WHO for developing the protocol which formed the basis for the study.

This research was solely funded by the Norwegian Institute of Public Health (NIPH) and received no specific grant from any funding agency, commercial or not-for-profit sectors.

