## Supplemental Files for "Household transmission of SARS-CoV-2; a prospective longitudinal study showing higher viral load and transmissibility of the Alpha variant compared to previous strains"

### Supplementary Methods

#### Definition of variables

Household size was defined as the number of people living in the household. Overcrowding was defined as 1) the number of rooms in the property being lower than the number of persons living in the household, AND 2) the number of square meters being less than 25 per person (1). Clinical severity was categorized as asymptomatic (no reported symptoms within 14 days after PCR positive test or reported "not feeling ill"), mild (reported symptoms except dyspnea, or reported feeling "almost not ill", "moderately ill" or "pretty ill"), or moderate (reported dyspnea or reported feeling "seriously ill"). Symptom onset was defined as the date of presence of either coughing, sore throat, runny nose, stuffy nose, dyspnea, fever, chills, change in taste/smell, headache, aches/pains, fatigue, nausea/vomiting, stomach pain/diarrhea, or a self-defined "date of symptom onset" (primary case only).

Duration of detectable SARS-CoV-2 was defined as the mean number of days a household contact was positive for laboratory confirmed SARS-CoV-2, i.e. the date of the last positive saliva test minus the date of the first positive saliva test according to the test regime (sampling at day 0, 3, 7, 10, 14, 21, 28 and 42). The date of the first positive sample was in some cases derived from the initial laboratory test taken through the municipality. Participants with any negative SARS-CoV-2 samples that had been diluted prior to RT-qPCR analysis were excluded from the analysis (n=7; 4 children and 3 adults), resulting in a total of 56 household contacts in the analysis (21 children and 35 adults).

#### Laboratory testing

##### *Detection of SARS-CoV-2 by RT-qPCR*

RNA was extracted from samples (200 µl) using MagNaPure 96 DNA and Viral NA Small Volume kits (no. 6543588001, Roche), and eluted in 50 ul. Saliva samples with too much mucus were mixed 1:1 with sputum lysis buffer containing N-acetylcystein (10 g/L) and shaken for 30 minutes. Viral transport medium was added to saliva samples with insufficient volume before extraction. Quantitative reverse transcription- polymerase chain reaction (RT-qPCR) was performed using the AgPath-ID One-step RT-

PCR kit (no. 4387391, Life Technologies) with primers and probes targeting two SARS-CoV-2 RdRp gene targets, developed at Institut Pasteur, Paris, France, and shared in the WHO protocol inventory (2). A 25 µl reaction was set up, containing 5 µl of RNA. Inconclusive results were resolved by repeating tests.

***Quantitative analysis of SARS-CoV-2 by droplet digital PCR (ddPCR)***

Results from the RT-qPCR analysis were evaluated to identify samples with high viral load that needed dilution to allow ddPCR quantification (3). The PCR reagents for the 2019-nCoV CDC ddPCR Triplex Probe Assay were assembled according to the manufacturer's instructions (Bio-Rad, Hercules, California, USA). Subsequent water/oil emulsion formation, PCR thermal cycling and final droplet reading in the QX200 Droplet Digital PCR system (Bio-Rad, Hercules, California, USA) were also done according to these instructions. The flow data were collected and initially analyzed by the QuantaSoft software (Bio-Rad) that accompanied the droplet reader. Final analysis of the ddPCR data in the Quantasoft Analysis Pro software (Bio-Rad) revealed the number of SARS-CoV-2 RNA copies per µl eluate. For saliva samples that were initially diluted due to insufficient volume, the dilution factor was taken into account for the estimation of the number of SARS-CoV-2 RNA copies per µl eluate.

***Sequencing of SARS-CoV-2***

Amplicon-based whole genome Sequencing (WGS) of SARS-CoV-2 was performed using the ARTIC-network nCoV-19 protocol v3 (4, 5) using either the Nanopore or Illumina (MiSeq) technology at the Norwegian Institute of Public Health (NIPH), or the Swift Amplicon SARS-CoV-2 Panel (Swift Bioscience) on Illumina (NovoSeq) at the Norwegian Sequencing Centre (NSC), according to the manufacturer's instructions with minor modifications. The pipelines used to generate consensus sequences are publicly available on the NIPH and NSC Github sites (6, 7). The phylogenetic assignment of the consensus sequences was performed using Pangolin (8).

***Blood typing***

Blood type was determined for all participants aged ≥18 years old using the Bio-Rad ID-Microtyping system at the Blood bank of Oslo University Hospital.

| | *Collected from all participants (n=216)* | *Collected only from participants aged ≥ 12 (n=166)* | |
|---|---|---|---|
| | **Saliva samples[a]** | **OP samples[a]** | **Blood samples** |
| **Day 0** | n= 198 (92%) | n= 156 (94%) | n= 151 (91%) |
| **Day 3** | n= 203 (94%) | n= 155 (93%) | |
| **Day 7** | n= 201 (93%) | n= 157 (95%) | n=147 (89%) |
| **Day 10** | n= 179 (83%) | n= 127(76%) | |
| **Day 14** | n= 191 (88%) | n= 147 (89%) | n=133 (80%) |
| **Day 21** | n= 170 (79%) | n= 123 (74%) | |
| **Day 28** | n= 160 (74%) | n= 127 (77%) | n=120 (72%) |
| **Day 42** | n= 156 (72%) | n= 113 (68%) | n=110 (66%) |

**Supplementary Figure S1:** Flow chart of sampling of the participants at the different timepoints throughout the study. The percentages indicate how many of the eligible participants at each timepoint who provided the different samples. Abbreviations; OP; oropharyngeal.

[a] collected for viral detection by PCR.

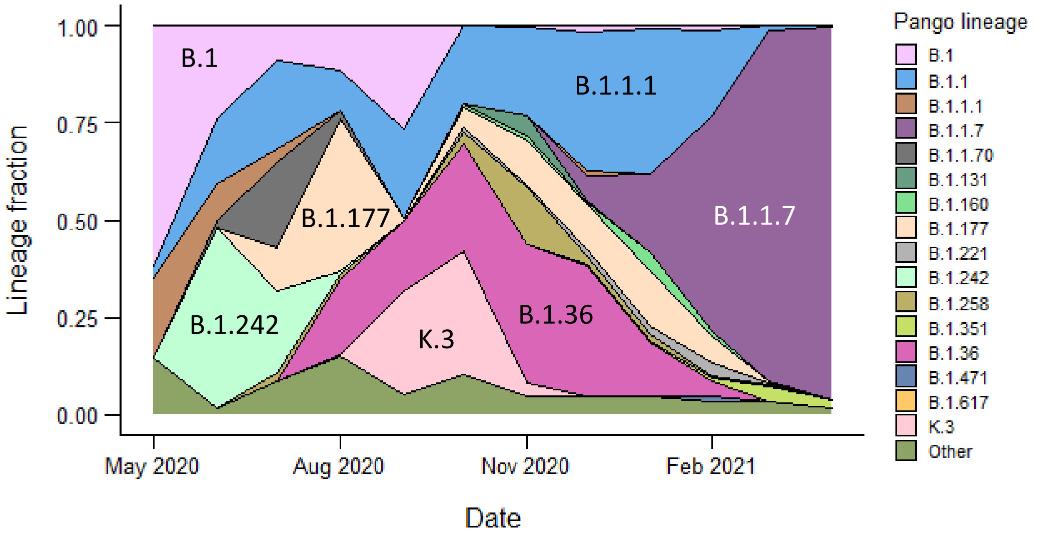


**Supplementary Figure S2:** The proportion of genetic subgroups of all SARS-CoV-2 viruses analyzed by Next Generation Sequencing (NGS) from Oslo and Viken, i.e. the counties of recruitment, per month during the study recruitment period, among sequences with >70% coverage. All subgroups with less than 5 occurrences are categorized as «Others», while «B» og «B.1» includes virus that were not allocated to a subgroup. Source: Surveillance data from the Norwegian Institute of Public Health (NIPH)

**Supplementary Table S1**: Frequency of participants with the various Pango lineages.

| Pango lineage | WHO label for VOCs[a] | Frequency |
|---|---|---|
| B.1.1.7 | Alpha | 46 |
| B.1.351 | Beta | 2 |
| B.1 | - | 2 |
| B.1.1.1 | - | 3 |
| B.1.1.141 | - | 1 |
| B.1.1.151 | - | 1 |
| B.1.1.153 | - | 1 |
| B.1.1.162 | - | 1 |
| B.1.1.277 | - | 6 |
| B.1.1.333 | - | 2 |
| B.1.1.39 | - | 1 |
| B.1.1.64 | - | 8 |
| B.1.36.21 | - | 16 |
| B.1.160 | - | 6 |
| B.1.177 | - | 10 |
| B.1.258 | - | 1 |
| B.1.367 | - | 7 |
| B.1.398 | - | 1 |
| K.3 | - | 7 |
| Missing | NA | 10 |

[a]VOCs; Variants Of Concern

**Supplementary Table S2:** Comparison of clinical severity according to genetic variant (N=123) or age (N=132) amongst confirmed cases

| Clinical severity | Genetic variant (n%[a]) | | | Age (n%[a]) | | |
|---|---|---|---|---|---|---|
| | **Alpha (N=46)** | **Non-VOC viruses (N=77)** | **p-value, chi2** | **2-17 yrs (N=31)** | **≥18 yrs (N=101)** | **p-value, chi2** |
| Asymptomatic | 10 (21.7) | 7 (9) | | 11 (35.5) | 8 (7.9) | |
| Mild | 16 (34.8) | 38 (49.4) | | 17 (54.8) | 40 (39.6) | |
| Moderate | 20 (43.5) | 32 (41.6) | 0.09 | 3 (9.7) | 53 (52.5) | **<0.00** |

Abbreviations: non-VOC; non- Variant Of Concern

[a] proportion of cases (%)

**Supplementary Table S3:** Comparison of clinical severity and symptoms according to genetic variant among primary cases only (N=58).

| **Clinical symptom** | **Alpha n (%[a]) (N=18)** | **Non-VOC viruses n (%[a]) (N=40)** | **p-value ($chi^2$)** |
|---|---|---|---|
| Severity | | | |
| Asymptomatic | 2 (11.1) | 3 (7.5) | |
| Mild | 5 (27.8) | 15 (37.5) | |
| Moderate | 11 (61.1) | 22 (55.0) | 0.74 |
| Cough | 16 (88.9) | 30 (75.0) | 0.23 |
| Fever | **15 (83.3)** | **18 (45.0)** | **0.01** |
| Dyspnea | 10 (55.6) | 21 (52.5) | 0.83 |
| Loss of taste/ smell | **16 (88.9)** | **23 (57.5)** | **0.02** |

Abbreviations: non-VOC; non variant of concern

[a] proportion of cases (%)

**Supplementary Table S4:** Association between ddPCR $log_{10}$ viral load (exposure) and symptoms (outcome).

| For all participants[a] | Crude OR (95% CI) | p-value | Adjusted[b] OR (95% CI) | p-value |
|---|---|---|---|---|
| **Symptom** | | | | |
| Loss of smell/taste | **1.39 (1.06-1.82)** | **0.02** | **1.40 (1.06-1.85)** | **0.02** |
| Cough | **1.46 (1.01-2.11)** | **0.045** | 1.37 (0.93-2.01) | 0.11 |
| Dyspnea | **1.36 (1.00-1.86)** | **0.048** | 1.34 (0.96-1.86) | 0.08 |
| Fever | 1.05 (0.82-1.35) | 0.69 | 1.04 (0.8-1.34) | 0.79 |
| **For primary cases[c]** | **Crude OR (95% CI)** | **p-value** | **Adjusted[b] OR (95% CI)** | **p-value** |
| **Symptom** | | | | |
| Loss of smell/taste | 1.43 (0.89-2.28) | 0.14 | 1.39 (0.85-2.28) | 0.19 |
| Cough | 1.15 (0.69-1.9) | 0.59 | 1.13 (0.68-1.89) | 0.64 |
| Dyspnea | 1.05 (0.71-1.56) | 0.80 | 1.06 (0.71-1.58) | 0.78 |
| Fever | 1.2 (0.8-1.8) | 0.38 | 1.2 (0.79-1.82) | 0.39 |

Abbreviations: OR; Odds Ratio, CI; Confidence Interval

[a] n=116, mixed-effect logistic regression model with a household-level random intercept

[b] adjusted for sex and age

[c] n=56, logistic regression
